# Knowledge, Attitudes and Practices (KAP) on Dengue Fever in Burkina Faso: findings from the national household survey in Burkina Faso

**DOI:** 10.64898/2026.02.11.26346064

**Authors:** Nicolas Ouédraogo, Siaka Débé, Harouna Soré, Farida Tiendrébégo, Gérard Wendyam Nonkani, Guillaume Sylvestre Sanou, Réné Kinda, Adama Ganou, Casimire Wendlamita Tarama, Sonia Ilboudo, Moussa Wamdaogo Guelbéogo, Isaie Meda, Adama Gansane

## Abstract

**Background:** Dengue fever is an emerging public health concern in Burkina Faso, with increasing outbreaks and data gaps in population awareness. This study assessed knowledge, attitudes, and practices (KAP) related to dengue fever in all health regions of the country.

**Methodology/Principal Findings:** A nationwide cross-sectional household survey was conducted in May 2022 using a stratified two-stage cluster sampling design. One rural and one urban area were selected per region. Heads of households or their representatives were interviewed using a structured questionnaire. Data were collected electronically and analyzed using Stata. A total of 1,568 participants were enrolled (52.0% male; 48.0% female). Overall, 66.3% had heard of dengue, with higher awareness in urban than rural areas. Only 49.0% correctly identified mosquito bites as the mode of transmission, and 29.9% did not know what dengue is. Most respondents (88.3%) stated that dengue can affect everyone. Regarding prevention, 80.2% reported sleeping under a mosquito net, 49.0% eliminated stagnant water, and 45.4% used mosquito repellents. In practice, 67.6% consistently slept under mosquito nets and 82.6% used repellents. Almost all respondents (98.6%) reported that they would consult a health professional if they had symptoms of dengue. However, knowledge about treatment and vaccination was limited, with 46.5% and 56.6% respectively reporting not knowing whether drugs or vaccines exist.

**Conclusions/Significance:** This study highlights moderate awareness but substantial knowledge gaps and urban–rural disparities in dengue-related KAP in Burkina Faso. Strengthening community-based education and integrated vector control strategies is essential to improve prevention and reduce dengue transmission.

**Author Summary:** Dengue fever is a mosquito-borne disease that is spreading in many parts of the world, including West Africa. In Burkina Faso, outbreaks have been reported in recent years, but little is known about what communities understand about the disease or how they protect themselves. In this study, we conducted a nationwide household survey covering all health regions of the country to explore people’s knowledge, perceptions, and prevention practices related to dengue.

We found that while many people had heard about dengue, important gaps remain. A significant proportion did not know how the disease is transmitted or whether treatments or vaccines exist. Preventive actions such as sleeping under mosquito nets and removing standing water were reported, but these practices were not consistent everywhere, especially between urban and rural areas. Encouragingly, almost all respondents said they would seek care from a health professional if they developed symptoms.

Our work provides the first national picture of community awareness and behaviors related to dengue in Burkina Faso. These findings highlight the need for strengthened health education and community engagement to improve prevention and support ongoing efforts to control mosquito-borne diseases.

## 1. Introduction

Dengue fever (DF) is a primary viral disease transmitted to humans by mosquitoes (*Aedes aegypti et Aedes albopictus)*, imposing a significant economic and health burden in numerous regions globally [1–3]. The global incidence of dengue has markedly increased over the past two decades, posing a substantial public health challenge. From 2000 to 2019, the World Health Organization documented a ten-fold surge in reported cases worldwide, increasing from 500,000 to 5.2 million. The year 2019 marked an unprecedented peak, with reported instances spreading across 129 countries. In 2023, dengue fever, has resurfaced in several endemic areas around the world, particularly in Burkina Faso, which is already experiencing a difficult security situation [4]. The first cases of dengue fever have been reported in Burkina Faso as early as 1925 and again in 1980 [5]. Larger outbreaks occurred in 2016 and 2017, leading to the strengthening of surveillance system in the country through the establishment of sentinel sites [6]. In 2016, the country recorded 2,600 cases and 21 deaths, followed by 14,944 cases and 30 deaths in 2017.

In 2023, the country becomes the most affected in the African region, experiencing a significant increase in dengue cases compared with the same periods in 2021 and 2022. outbreak in 2023 showed higher trends in mortality and morbidity compared to those seen in 2016 and 2017with a cumulative number of cases reported in the country for 2023 is 146,878 suspected cases, including 68,346 probable cases (positive rapid diagnostic test) and 688 deaths among suspected cases, representing a case fatality rate of 0.5% [4,7].

Despite the efforts of the Ministry of Health, many people in Burkina Faso have turned to alternative treatments, including traditional medicine very often inappropriate or badly dosed, to treat or prevent dengue fever [8]. A major challenge in controlling the disease is the confusion between dengue fever and malaria, as both diseases present with similar symptoms such as fever, headache, and body aches [9]. This misidentification leads to delays in seeking appropriate medical care and contributes to the spread and severity of the disease. Furthermore, the proliferation of unverified treatment methods and misinformation about the disease complicates public health efforts to control the outbreak[8].

Understanding the knowledge, attitudes, and practices (KAP) of the population regarding dengue fever is crucial for designing effective public health interventions. Assessing the level of awareness and behavior of the population will help identify gaps in knowledge and guide the development of targeted strategies for dengue prevention and treatment. The aim of this study was to assess the population’s knowledge, attitudes and practices relating to dengue fever in the 13 health regions of Burkina Faso by means of a survey of the population (urban and rural).

## 2. Methods

### 2.1. Study sites

The study was carried out throughout the country, i.e. in the 13 regions of the country. In each region, one rural site and one urban site in the regional capital were selected. The study sites are described in Figure 1.

**Figure 1:**
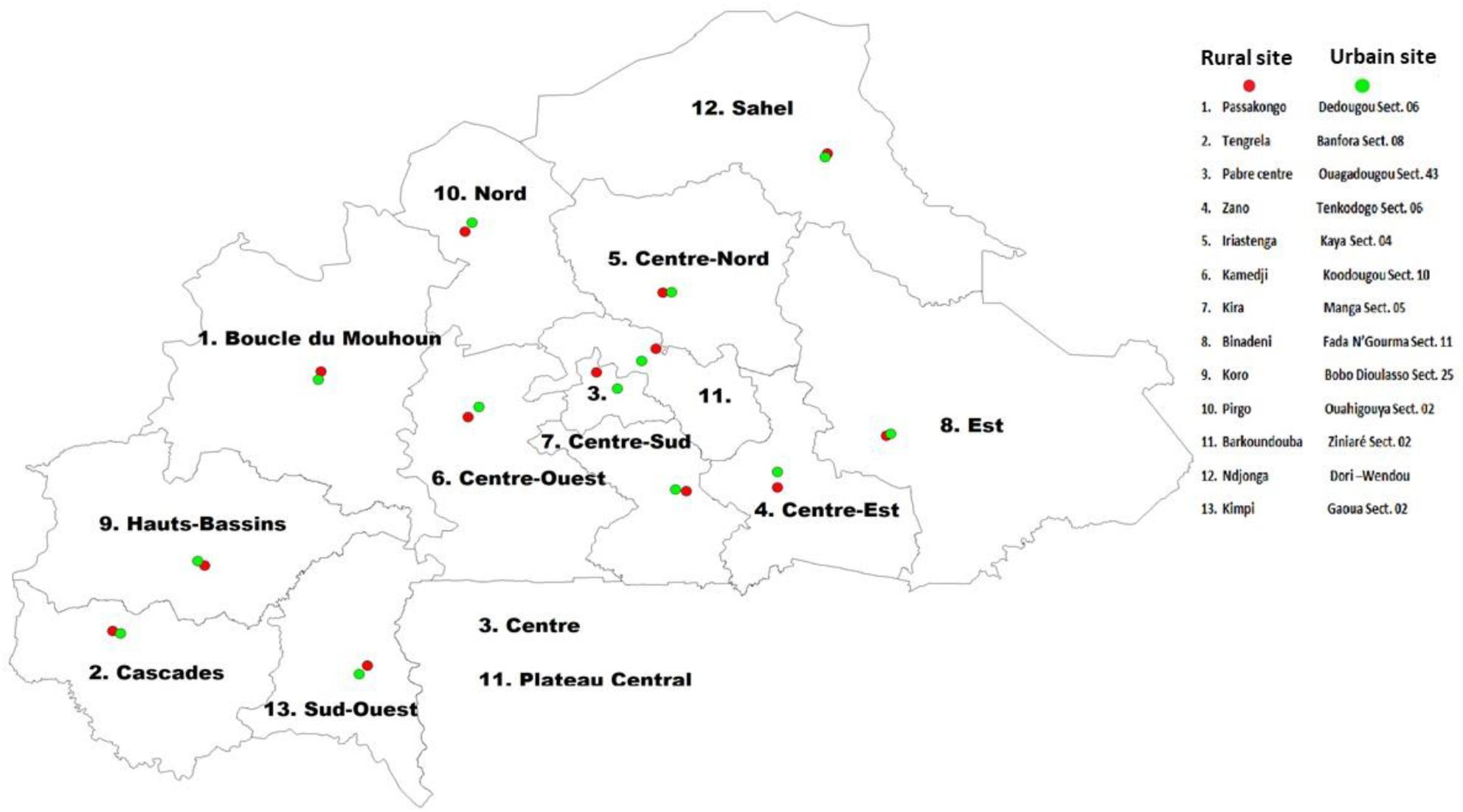
study sites.

### 2.2. Participant selection method

A sampling plan combining stratified and cluster sampling was used. Stratification was based on the country’s 13 regions, and in each of these regions on the rural (peri-urban) and urban areas of each capital. Cluster sampling was based on the sampling frame available from the 2019 general population and housing census (GPHC 2019), and the sample was selected at two levels: enumeration areas and households.

#### 2.2.1. First stage: choice of enumeration area

The enumeration areas (EA) of the GPHC were used as clusters. Each EA comprises at least 250 households. In each stratum, 01 EA was selected. This choice was made by simple random selection.

#### 2.2.2. Second stage: selection of households

Households were selected on the basis of a systematic random sampling plan in each of the enumeration areas selected. When a selected household was not eligible, it was replaced by another household on the waiting list in chronological order.

#### 2.2.3. Selection and administration of the participant questionnaire in households

In each selected household, the head was invited to take part in the cross-sectional study using the questionnaire in appendix.

### 2.3. Data Collection

The questionnaire consisted of two sections: one covering the participants’ socio-demographic characteristics, and the other assessing their knowledge, attitudes, and practices (KAP) related to dengue. The KAP survey was conducted in selected households as part of a cross-sectional screening study in May 2022. Data were collected using a previously validated questionnaire, developed based on a review of published KAP studies and questionnaires provided by their authors. Information was gathered through individual interviews with either the head of the household or a designated representative. The questionnaire was administered in French or, when necessary, in the participant’s local language with the assistance of a local interpreter, who also served as a guide to facilitate household access.

### 2.4. Data Management and analysis

The study data were entered directly into an electronic CRF configured on tablets. The database was then extracted in Excel format. Data analysis was carried out using Stata software. DIVA-GIS software was used to determine the study areas.

### 2.5. Ethics statement

The project protocol received approval from the National Ethics Committee prior to the implementation of project activities under reference No. 2020-9-195, dated September 2, 2020. Authorization was also obtained from the administrative, health, and security authorities in each region. Before data collection, informed consent was obtained from household heads or their representatives for household-level data, and from each individual participant for personal data collection. Participation was entirely voluntary.

## 3. Results

This study, conducted across all 13 health regions of Burkina Faso with a balanced inclusion of urban and rural populations, provides critical insights into the knowledge, attitudes, and practices (KAP) of the population regarding dengue fever. The findings reveal both promising awareness levels and significant gaps that warrant targeted public health interventions.

### 3.1. Socio-Demographic Patterns

A total of 1,568 participants were enrolled in the study across 26 sites nationwide. In each of the 13 health regions of Burkina Faso, one rural and one urban site were selected, ensuring balanced geographic and demographic representation. The distribution of participants by region and site is presented in Figure 2.

**Figure 2:**
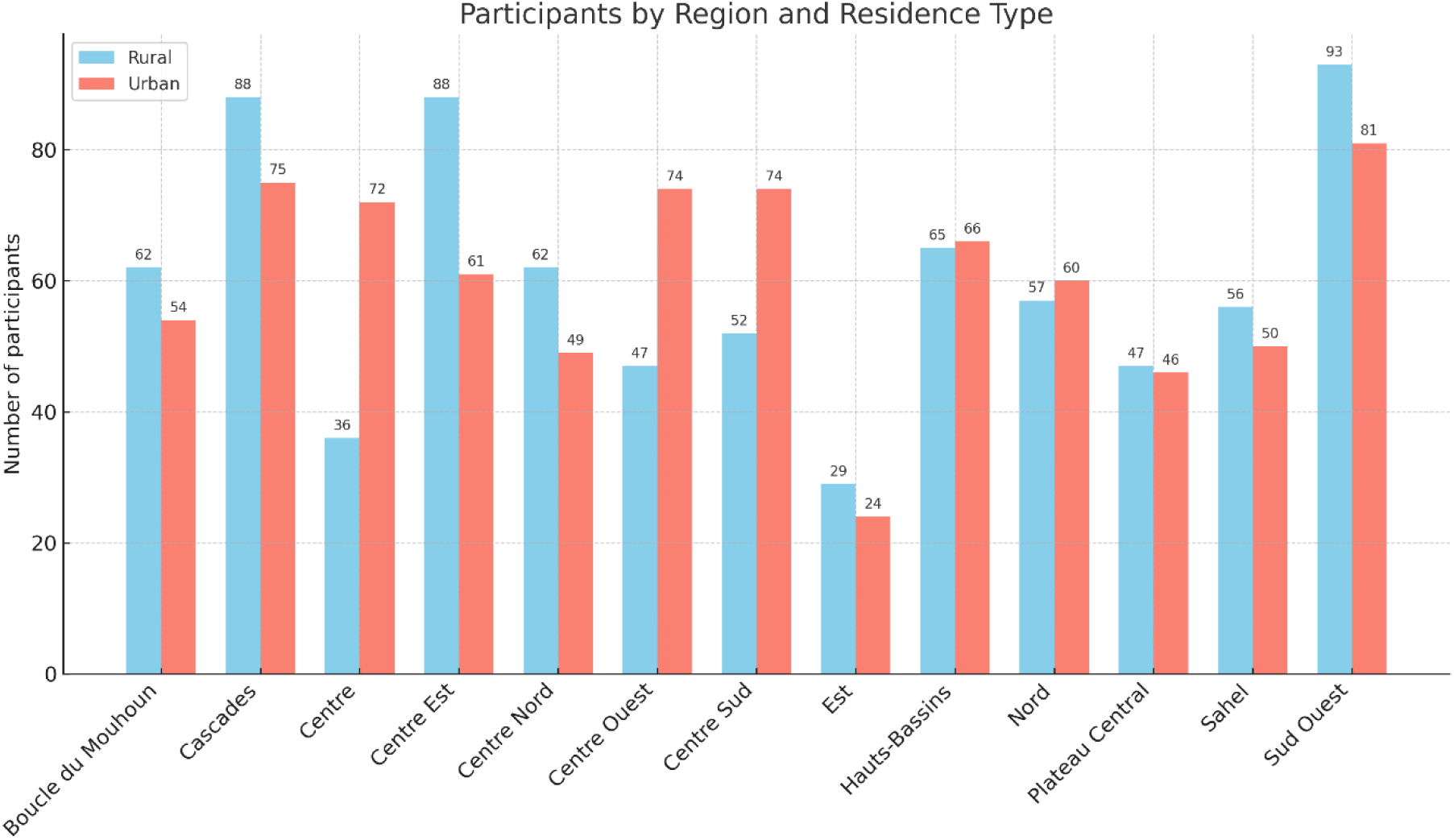
Participant’s distribution by region and site.

After obtaining informed consent from the participants, their socio-demographic characteristics were collected. These characteristics are presented in Table 1. Among the 1,568 respondents, 52.04% were male and 47.96% female, with a nearly equal distribution between rural and urban areas. The most represented age groups were 35–44 years (24.11%), 25–34 years (21.17%), and 45–54 years (20.15%). Most participants were married in monogamy (57.33%) or polygamy (23.22%). Over half (54.70%) had no formal schooling, and farming was the main occupation (38.26%), followed by trading (13.31%) and unemployment (12.41%).

**Table 1:** Socio-Demographic characteristics of study population (n=1568)

| Variables | Respondents distribution |  |  |  |  |  |
| --- | --- | --- | --- | --- | --- | --- |
|  | Rural |  | Urban |  | Total |  |
|  | n=782 | % | n= 786 | % | n=1568 | % |
| **Sex:** |  |  |  |  |  |  |
| Male | 413 | 26.34 | 403 | 25.7 | 816 | 52.04 |
| Female | 369 | 23.53 | 383 | 24.43 | 752 | 47.96 |
| **Age:** |  |  |  |  |  |  |
| 15-24 | 77 | 4.91 | 84 | 5.36 | 161 | 10.27 |
| 25-34 | 146 | 9.31 | 186 | 11.86 | 332 | 21.17 |
| 35-44 | 185 | 11.80 | 193 | 12.31 | 378 | 24.11 |
| 45-54 | 168 | 10.71 | 148 | 9.44 | 316 | 20.15 |
| 55-64 | 115 | 7.33 | 80 | 5.10 | 195 | 12.44 |
| 65-74 | 60 | 3.83 | 65 | 4.15 | 125 | 7.97 |
| 75-84 | 26 | 1.66 | 28 | 1.79 | 54 | 3.44 |
| 85-94 | 3 | 0.19 | 2 | 0.13 | 5 | 0.32 |
| 95+ | 2 | 0.13 | 0 | 0.00 | 2 | 0.13 |
| **Marital satatus:** |  |  |  |  |  |  |
| Single | 42 | 2.69 | 102 | 6.53 | 144 | 9.21 |
| Divorcee | 0 | 0 | 2 | 0.13 | 2 | 0.13 |
| Married in monogamy | 412 | 26.36 | 484 | 30.97 | 896 | 57.33 |
| Married in polygamy | 242 | 15.48 | 121 | 7.74 | 363 | 23.22 |
| Widow/widower | 8 | 5.12 | 70 | 4.48 | 150 | 9.6 |
| NA | 2 | 0.13 | 6 | 0.38 | 8 | 0.51 |
| **Level of education:** |  |  |  |  |  |  |
| Literacy | 27 | 1.73 | 99 | 6.33 | 126 | 8.06 |
| No schooling | 541 | 34.61 | 314 | 20.09 | 855 | 54.7 |
| Primary level | 85 | 5.44 | 98 | 6.27 | 183 | 11.71 |
| Secondary level | 54 | 3.45 | 197 | 12.6 | 251 | 16.06 |
| University level | 2 | 0.13 | 76 | 4.86 | 78 | 4.99 |
| NA | 69 | 4.41 | 1 | 0.06 | 70 | 4.48 |
| **Occupation:** |  |  |  |  |  |  |
| Trader | 27 | 1.73 | 181 | 11.58 | 208 | 13.31 |
| Farmer | 503 | 32.18 | 95 | 6.08 | 595 | 38.26 |
| Breeder | 36 | 2.3 | 6 | 0.38 | 42 | 2.69 |
| Housewife | 96 | 6.15 | 12 | 0.77 | 108 | 6.92 |
| Pupil/ Student | 19 | 1.22 | 52 | 3.33 | 71 | 4.54 |
| Unemployed | 51 | 3.26 | 143 | 9.15 | 194 | 12.41 |
| Private sector | 14 | 0.9 | 87 | 5.57 | 101 | 6.46 |
| Retired | 1 | 0.06 | 18 | 1.15 | 19 | 1.22 |
| Public sector | 3 | 0.19 | 109 | 6.97 | 112 | 7.17 |
| Other | 11 | 0.7 | 80 | 5.12 | 91 | 5.82 |
| NA | 17 | 1.09 | 2 | 0.13 | 19 | 1.22 |

### 3.2. Knowledge of Dengue Fever

In order to assess the KAP on dengue fever, an initial general question was asked to all participants: Are you aware of the existence of a disease called Dengue? The answers to this question are set out in the Table 2. This first question allows us to exclude all those whose answer is no and to continue our survey with those whose answer is yes.

**Table 2:** Distribution of respondents aware of the existence of dengue fever, by place of residence (n = 1,568)

| Response | Rural (n, %) | Urban (n, %) | Total (n, %) |
| --- | --- | --- | --- |
| Yes | 419 (26.72%) | 621 (39.60%) | 1,040 (66.33%) |
| No | 363 (23.15%) | 165 (10.52%) | 528 (33.67%) |
| Total | 782 (49.87%) | 786 (50.13%) | 1,568 (100%) |

Analysis of household sources of information on dengue fever in Burkina Faso reveals a diversity of channels used by the population to access information. Knowledge of these sources is vital if we are to raise awareness and combat dengue fever more effectively. Figure 3 is a radar diagram illustrating the proportions of information channels on dengue fever used by participants in the cross-sectional survey of the present study.

**Figure 3:**
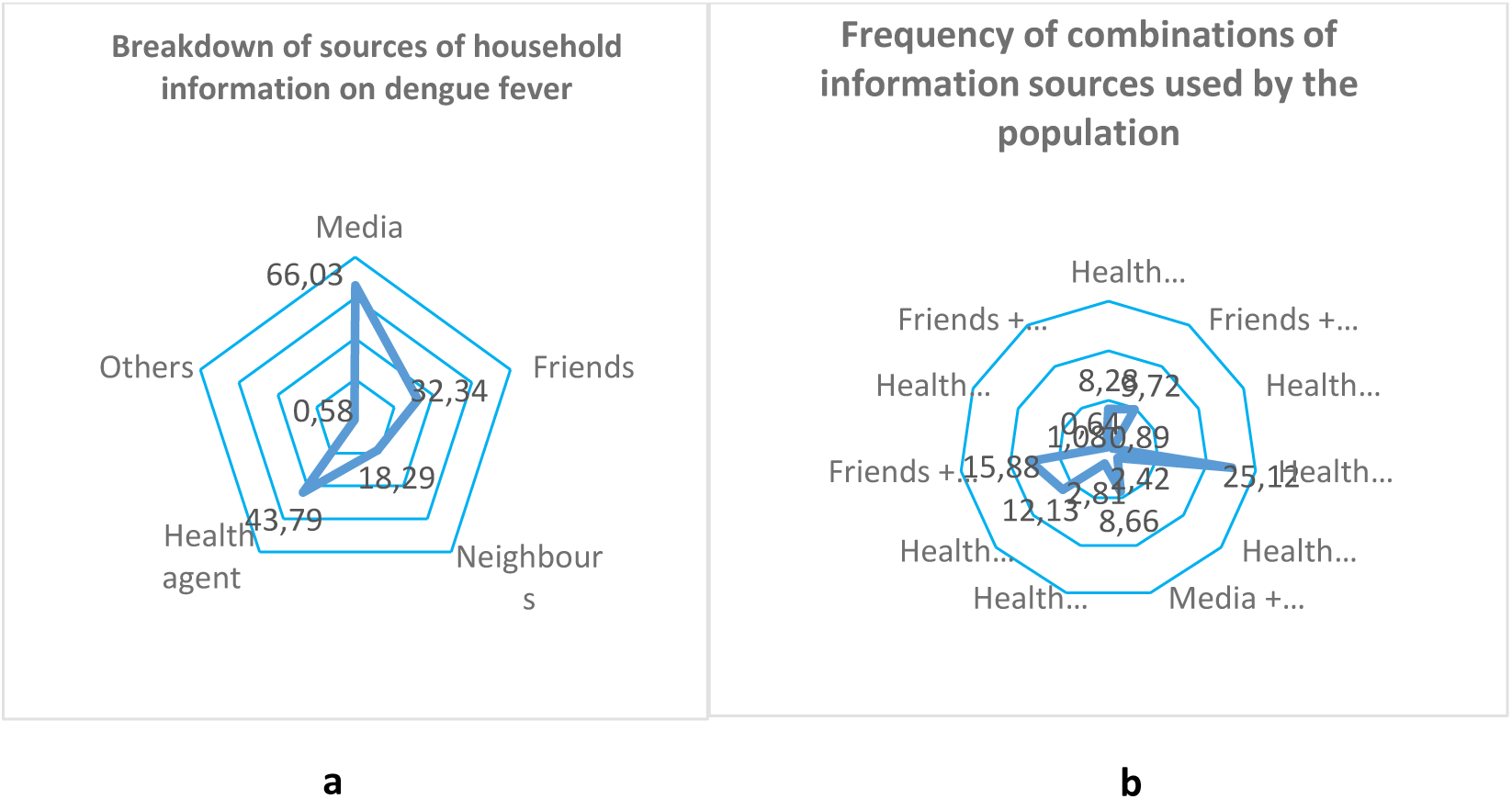
Source of information on dengue fever among participants.

Table 3 provides insights into participants’ knowledge regarding symptoms of Dengue fever virus infection.

**Table 3:**
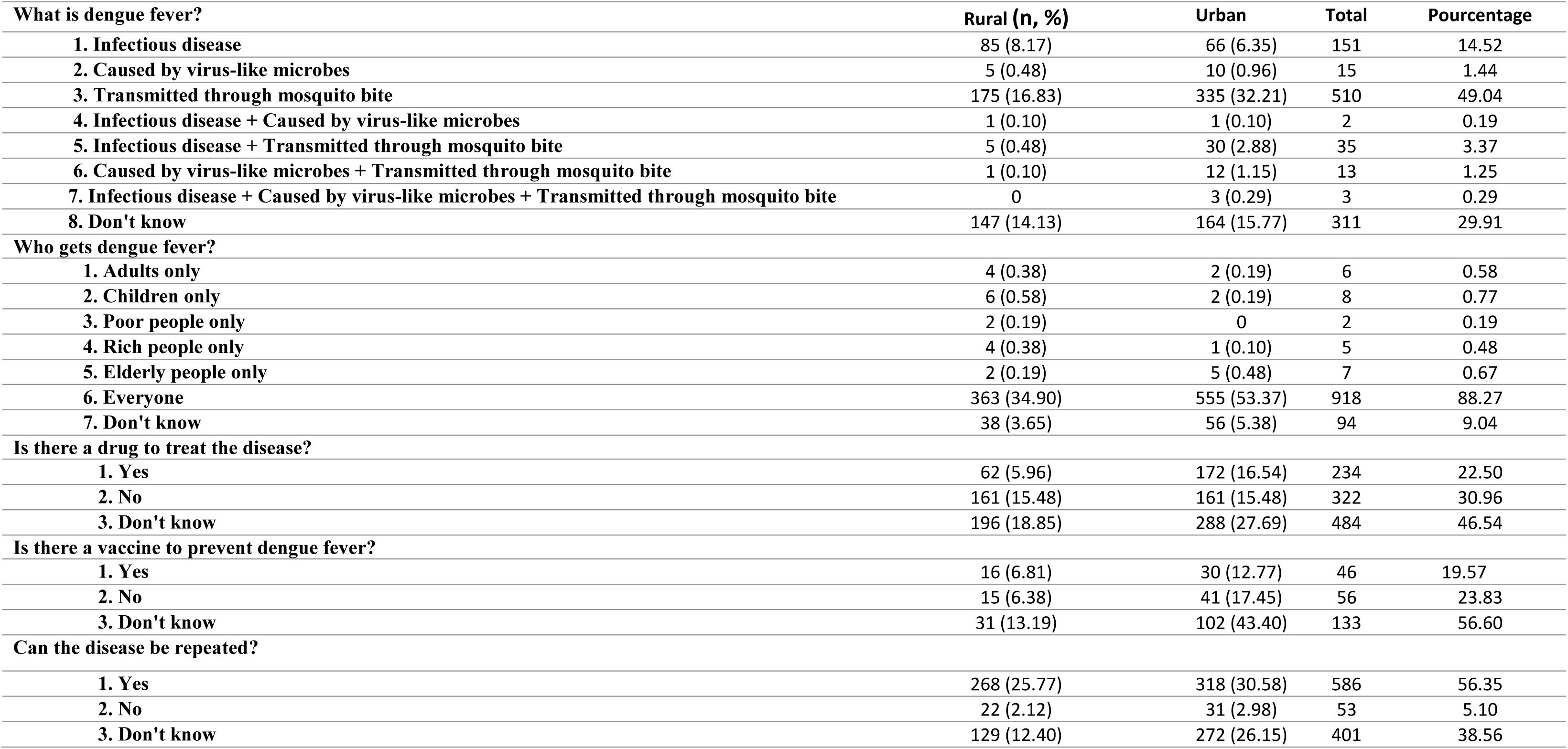
Knowledge towards dengue fever.

Overall, 49.04% of respondents identified dengue as a disease transmitted through mosquito bites, while 14.52% described it as an infectious disease and 29.91% reported that they did not know what dengue is. Only 1.44% mentioned that dengue is caused by virus-like microbes, and 0.29% correctly combined all three aspects (infectious, viral, and mosquito-borne).

Regarding populations affected, 88.27% stated that dengue can affect everyone, whereas 9.04% did not know. Very small proportions indicated that dengue affects only adults (0.58%), children (0.77%), the poor (0.19%), the rich (0.48%), or the elderly (0.67%).

Concerning treatment, 22.50% of respondents reported that a drug exists to treat dengue, 30.96% said no, and 46.54% did not know.

With respect to prevention, 19.57% reported the existence of a vaccine against dengue, 23.83% stated that there is no vaccine, and 56.60% did not know.

Finally, 56.35% indicated that dengue can be repeated, 5.10% said it cannot be repeated, and 38.56% did not know.

### 3.3. Attitudes Towards Dengue Prevention and Treatment

The measures taken by study participants to protect themselves against dengue fever, treat themselves if they contract the disease, or treat a relative who is ill with dengue fever are listed in Table 4. Regarding personal protection, 80.19% of respondents reported sleeping under a mosquito net, 49.04% reported eliminating stagnant water, and 45.38% reported using mosquito repellent. Only 22.21% reported wearing long, loose clothing, while 7.12% stated that they do nothing to protect themselves from dengue. Other preventive actions such as cleaning the living environment were rarely mentioned (0.48%).

**Table 4:** Attitudes towards Dengue Fever.

| What do you do to protect yourself from dengue fever? |  | Rural | Urban | Total | Pourcentage |
| --- | --- | --- | --- | --- | --- |
| **1. Wear long, loose clothing** |  |  |  |  |  |
|  | No | 283 (27.21) | 526 (50.58) | 809 | 77.79 |
|  | Yes | 136 (13.08) | 95(9.13) | 231 | 22.21 |
| **2. Use mosquito repellent** |  |  |  |  |  |
|  | No | 216(20.77) | 352(33.85) | 568 | 54.62 |
|  | Yes | 203(19.52) | 269(25.87) | 472 | 45.38 |
| **3. Eliminate stagnant water** |  |  |  |  |  |
|  | No | 234(22.50) | 296(28.46) | 530 | 50.96 |
|  | Yes | 185(17.79) | 325(31.25) | 510 | 49.04 |
| **4. Sleep under a mosquito net** |  |  |  |  |  |
|  | No | 51(4.90) | 155(14.90) | 206 | 19.81 |
|  | Yes | 368(35.38) | 466(44.81) | 834 | 80.19 |
| **5. Other (specify): Don't know and clean up the living environment** |  |  |  |  |  |
|  | No | 417(40.10) | 618(59.42) | 1035 | 99.52 |
|  | Yes | 2(0.19) | 3(0.29) | 5 | 0.48 |
| **6. Do nothing** |  |  |  |  |  |
|  | No | 399(38.37) | 567(54.52) | 966 | 92.88 |
|  | Yes | 20(1.92) | 54(5.19) | 74 | 7.12 |
| **If you think you have dengue fever and you have symptoms, what do you do?** |  |  |  |  |  |
| **1. Consult a doctor** |  |  |  |  |  |
|  | No | 8(0.77) | 7(0.67) | 15 | 1.44 |
|  | Yes | 411(39.52) | 614(59.04) | 1025 | 98.56 |
| **2. Self-medication** |  |  |  |  |  |
|  | No | 410(39.42) | 617(59.33) | 1027 | 98.75 |
|  | Yes | 9(0.87) | 4(0.38) | 13 | 1.25 |
| **3. Use of plants** |  |  |  |  |  |
|  | No | 375(36.06) | 604(58.08) | 979 | 94.13 |
|  | Yes | 44(4.23) | 17(1.63) | 61 | 5.87 |
| **4. Consult a traditional healer** |  |  |  |  |  |
|  | No | 416(40) | 620(59.62) | 1036 | 99.62 |
|  | Yes | 3(.29) | 1(0.10) | 4 | 0.38 |
| **5. Prayer** |  |  |  |  |  |
|  | No | 414(39.81) | 604(58.08) | 1018 | 97.88 |
|  | Yes | 5(0.48) | 17(1.63) | 22 | 2.12 |
| **6. Other (specify)** |  |  |  |  |  |
|  | No | 418(40.19) | 621(59.71) | 1039 | 99.90 |
|  | Yes | 1(0.10) | 0(0.0) | 0 | 0.10 |
| **7. Do nothing** |  |  |  |  |  |
|  | No | 418(40.19) | 615(59.13) | 1033 | 99.33 |
|  | Yes | 1(0.10) | 6(0.58) | 7 | 0.67 |
| **If you suspect someone has dengue fever, what do you do?** |  |  |  |  |  |
| **1. Advise them to go to a health Centre** |  | 411 (39.52) | 615 (59.13) | 1026 | 98.65 |
| **2. Approach them to go to a traditional practitioner** |  | 1 (0.10) | 0 | 1 | 0.10 |
| **3. Approach them to start self-medicating** |  | 4 (0.38) | 0 (0.00) | 4 | 0.38 |
| **4. Do Nothing** |  | 3 (0.29) | 6 (0.58) | 9 | 0.87 |

When experiencing symptoms suggestive of dengue, 98.56% of respondents reported that they would consult a doctor. Very few indicated self-medication (1.25%), use of medicinal plants (5.87%), consultation of a traditional healer (0.38%), prayer (2.12%), or doing nothing (0.67%). If they suspected someone else had dengue, 98.65% stated that they would advise the person to go to a health centre. Only small proportions reported advising traditional treatment (0.10%), self-medication (0.38%), or doing nothing (0.87%).

### 3.4. Household Practices towards Dengue Prevention and Treatment

Participants’ practices for dengue prevention and treatment are recorded in Table 5 below. Overall, 67.60% of respondents reported sleeping under a mosquito net all the time, while 32.40% did not. Only 8.46% of households had mosquito netting on windows and 3.65% had netting on doors.

**Table 5:** Practices towards dengue fever.

| Dengue practices | Rural | Urban | Total | Pourcentage |
| --- | --- | --- | --- | --- |
| **Do you sleep under a mosquito net all the time?** |  |  |  |  |
| No | 126(12.12) | 211(20.29) | 337 | 32.40 |
| Yes | 293(28.17) | 410(39.42) | 703 | 67.60 |
| **Does your household have mosquito netting on the windows?** |  |  |  |  |
| No | 410(39.42) | 542(52.12) | 952 | 91.54 |
| Yes | 9(0.87) | 79(7.60) | 88 | 8.46 |
| **Does your household have mosquito netting on the door of the main house?** |  |  |  |  |
| No | 413(39.71) | 589(56.63) | 1002 | 96.35 |
| Yes | 6(0.58) | 32(3.08) | 38 | 3.65 |
| **Other methods used to control mosquitoes:** |  |  |  |  |
| 1. Mosquito repellent spirals and sprays |  |  |  |  |
| No | 153(14.71) | 28(2.69) | 181 | 17.40 |
| Yes | 266(25.58) | 593(57.02) | 859 | 82.60 |
| 2. Use of repellent herbs or plants |  |  |  |  |
| No | 273(26.25) | 579(55.67) | 852 | 81.92 |
| Yes | 146(14.04) | 42(4.04) | 188 | 18.08 |
| 3. Other (please specify):none, mosquito net, environmental hygiene, don't know, property, clean up your living environment, mosquito repellent, anti-mosquito ointment, turn the fan on all night, close the windows of houses every evening, avoid storing stagnant water, clean up your surroundings |  |  |  |  |
| No | 387(37.21) | 603(57.98) | 990 | 95.19 |
| Yes | 32(3.08) | 18(1.73) | 50 | 4.81 |
| **What medication do you use if you notice signs of dengue fever?** |  |  |  |  |
| 1. Modern medicine | 414(39.81) | 610(58.65) | 1024 | 98.46 |
| 2. Traditional medicine | 2(0.19) | 10(0.96) | 12 | 1.15 |
| 3. Other (please specify): Don't know | 3(0.29) | 1(0.10) | 4 | 0.38 |

Most respondents (82.60%) reported using mosquito repellent spirals or sprays to control mosquitoes, whereas 18.08% used repellent herbs or plants and 4.81% reported other methods such as environmental hygiene or eliminating stagnant water.

Regarding treatment practices, 98.46% stated that they use modern medicine when they notice signs of dengue fever, while 1.15% reported using traditional medicine and 0.38% did not know.

## 4. Discussion

Of the 1,568 participants; 782 were recruited from rural sites and 786 from urban sites. Urban residents were more numerous in the Centre, Cascades, and Centre Est regions, whereas rural residents predominated in Boucle du Mouhoun, Sud-Ouest, Est, and Sahel. The Centre Sud region had an equal number of participants from both settings. This distribution supports regionally and residentially disaggregated analyses of knowledge, attitudes, and practices related to dengue. This demonstrates the quality of the information collected, given the balance between rural and urban areas, as well as national representativeness.

The study included 1,568 participants, with a nearly equal distribution of males (52.04%) and females (47.96%) across rural and urban areas. Most respondents were adults aged 25–54 years (65.43%), reflecting an active population likely to be exposed to dengue vectors. Similar results on the sex ratio and age range were obtained in other KAP studies among patients attending Health Facilities in Lagos, Nigeria and on health care workers in Somalia during a cross-sectional study [10,11]. Over half of the participants (54.7%) had no formal education, especially in rural areas, while higher education levels were more common in urban settings. This educational disparity may impact awareness and prevention practices related to dengue. The majority were married (80.55%), with polygamy more prevalent in rural areas. Farming was the main occupation (38.26%), particularly in rural zones, whereas urban respondents were more engaged in trade and public/private sectors. Unemployment was higher in urban areas. These socio-demographic differences highlight the need for targeted dengue control strategies, especially in rural areas with lower education levels and high exposure risk due to outdoor occupations.

Analysis of basic knowledge about dengue showed that 66.33% of respondents (n = 1040) said they were aware of the disease, while 33.67% (n = 528) had never heard of it.

There was a marked difference according to place of residence. In urban areas, 621 people (39.60%) answered in the affirmative, compared with 419 people (26.72%) in rural areas. Conversely, lack of awareness was more common in rural areas (23.15%) than in urban areas (10.52%). So, although the numbers are broadly balanced between rural (49.87%) and urban (50.13%) areas, urban residents appear to be significantly better informed about dengue fever than those in rural areas. This could be explained by better access to the media, closer proximity to health facilities, or a greater intensity of awareness campaigns in urban areas. Similar results were found in other countries [12,13]. These results highlight a gap in access to health information depending on where people live, which raises major concerns in terms of equity in prevention. It is essential to adapt public health communication strategies to boost awareness in rural areas, by mobilizing community relays, local health workers or targeted campaigns. Figure 3a shows the proportions of individuals informed according to five main sources identified: the media, health workers, friends, neighbours and other minor sources. It can be seen that the media is the most cited source (66.03%), followed by health workers (43.79%), friends (32.34%) and neighbours (18.29%), and marginally by other sources (0.58%). These results confirm that the media and health workers are the pillars of health communication within households, and show the moderate effect of close social networks in disseminating health messages. They are consistent with the conclusions of previous studies conducted in the sub-region, which identify the media and health professionals as the preferred vectors for raising awareness during dengue epidemics [14]. Saghir and col. in Seiyun, Hadramout Yemen, found that the media ranked as the source of information regarding dengue fever with a rate lower than ours at 24.80% [15]. Figure 3b takes this analysis further by illustrating the different combinations of information sources used. The most frequently reported combination is “health workers + media” (25.12%), which confirms the complementary nature of institutional information and media dissemination. This was followed by “friends + media” (15.88%), “health workers + friends” (12.13%), and “health workers + friends + media” (8.66%), reflecting the importance of informal exchanges as a relay for official campaigns. Certain combinations including four sources are very poorly represented, as are those involving neighbours or the “other” category, suggesting a selective and hierarchical use of information channels. Overall, more than half of the participants had been exposed to at least two sources of information, which may be a factor favouring the consolidation of knowledge and the adoption of preventive behavior [16,17]. These results show that, in the context of the fight against vector-borne diseases such as dengue, there is a need to explore other opportunities for diversifying information vectors, and to devise coherent, multi-source communication strategies combining the mass media, health professionals and interpersonal networks, in order to boost dengue awareness and equity of access to health information, particularly in rural areas where the media may be less accessible [16,17]. In short, this analysis underlines the strategic role of the media and health workers in the fight against dengue fever in Burkina Faso, while highlighting the need to strengthen community networks as complementary information relays. In addition, it suggests that exposure to several sources is associated with better dissemination of prevention messages, reinforcing the value of an integrated public health communication approach adapted to the national socio-cultural context.

The results of Table 3 showed varied levels of knowledge regarding dengue fever among respondents, with notable differences between rural and urban populations. Awareness of dengue fever remains limited. Only 14.52% of participants correctly identified dengue as an infectious disease, 49.04% recognized its transmission through mosquito bites and very few (1.44%) understood the viral etiology. Urban respondents were more likely to associate dengue with mosquito bites (32.21%) compared to rural respondents (16.83%). However, nearly 30% of all participants stated they didn’t know what dengue fever is, reflecting significant gaps in basic knowledge, particularly in rural areas. This finding aligns with prior studies conducted in sub-Saharan Africa, where lack of formal education and limited access to reliable information sources have been linked to poor disease awareness[10]. While 88.27%, the vast majority of respondents correctly acknowledged that dengue can affect everyone with a higher proportion of correct responses in urban areas (53.37%) than in rural areas (34.90%), misconceptions such as dengue affecting only children or the poor were still present in a minority, indicating relatively good awareness on disease susceptibility. This reinforces the importance of targeted public education to address persistent myths [18]. Knowledge about treatment and prevention was also inadequate. Only 22.5% believed there is a drug to treat dengue, with urban participants (16.54%) being more likely to say yes than rural participants (5.96%), while 46.54% did not know. Similarly, more than half (56.60%) were unaware of the existence of a vaccine, reflecting a missed opportunity to promote immunization, particularly in high-risk areas. This uncertainty reflects either a lack of accurate information or confusion between supportive care and curative treatment. A majority of participants (56.35%) correctly stated that dengue can occur more than once. This awareness was higher among urban respondents (30.58%) compared to rural ones (25.77%). However, over one-third (38.56%) still answered “don’t know,” indicating the need to strengthen knowledge about disease recurrence and immunity. These findings demonstrate that urban populations generally have greater knowledge about dengue fever than rural populations, particularly regarding transmission, reinfection, and treatment. However, misinformation and lack of awareness remain widespread in both settings. Targeted health education campaign-especially in rural are essential to improve understanding of dengue prevention and control.

Results about attitude toward Dengue fever in Table 4 showed that the use of mosquito nets emerged as the main preventive strategy, reported by 80.19% of respondents, with a slightly higher proportion in urban areas (44.81%) compared to rural areas (35.38%). This difference was statistically significant (χ² = 24.96, p < 0.001). Only 22.21% of participants reported wearing long, loose clothing as protection against mosquito bites, with rural residents (13.08%) significantly more likely to adopt this measure than urban residents (9.13%) (χ² = 41.65, p < 0.001). Mosquito repellent use was reported by 45.38% of participants, with slightly more respondents in urban settings (25.87%) than rural areas (19.52%), although the difference was not statistically significant. Eliminating stagnant water, a key environmental control measure, was practiced by 49.04% of participants, significantly more in urban (31.25%) than in rural areas (17.79%) (χ² = 6.38, p = 0.011). Only 0.48% cited “other measures” (e.g., cleaning the living environment or not knowing what to do). Doing nothing was reported by 7.12% overall, with a slightly higher prevalence in urban areas (5.19%) than rural areas (1.92%) (χ² = 5.25, p = 0.022). In summary, while some preventive measures, notably bed net use, are widely adopted, others such as repellent use, protective clothing, and source reduction are less frequently practiced. Significant rural and urban differences suggest that contextual factors such as access to resources, housing types, and exposure risks influence the choice of prevention strategies.

When asked what they would do if they suspected having dengue fever and experienced symptoms, the vast majority of respondents (98.56%) reported that they would consult a physician, with no significant difference between rural and urban areas. Other responses were far less frequent: self-medication (1.25%), use of medicinal plants (5.87%), consulting a traditional healer (0.38%), and prayer (2.12%). The use of plants was significantly more common in rural areas (4.23%) than in urban areas (1.63%) (χ² = 25.92, p < 0.001). Only 0.67% indicated they would “do nothing” if symptomatic, and differences between rural and urban areas were not statistically significant. These findings point to a strong preference for formal healthcare services over traditional or alternative methods, reflecting generally favorable health-seeking behavior in both rural and urban populations. When asked what they would do if they suspected someone had dengue fever, 98.65% of respondents reported that they would advise the individual to seek care at a health centre, with no significant differences between rural and urban areas. Approaching the person for traditional practitioners (0.10%) or self-medication (0.38%) were rare, and doing nothing was reported by less than 1% (0.87%). This strong tendency to refer suspected cases to healthcare facilities demonstrates high awareness of the importance of early medical care, a positive element for community-based dengue control. The data reveal high awareness and positive attitudes towards medical consultation in both rural and urban contexts. This finding agrees with results elsewhere, including Sudan (74.1%), Northern Iran (81%), and Eastern Ethiopia (52%)[19–21]. However, there is a marked discrepancy in the adoption of some preventive measures. Rural participants are more likely to rely on protective clothing and plants, whereas urban residents are more engaged in environmental management (eliminating stagnant water) and slightly more likely to do nothing. These differences may reflect disparities in resource availability, housing structures, urban infrastructure, and exposure patterns. Public health interventions should emphasize integrated preventive strategies; combining personal protection and environmental management while tailoring messages to the local context. Leveraging the widespread use of mosquito nets, already promoted through malaria control campaigns, could be an effective entry point for reinforcing dengue prevention messages. Mosquito control is critical, comprehensive dengue prevention also involves community engagement, environmental management, early diagnosis, and public awareness campaigns.

The majority of respondents (67.6%) reported consistently sleeping under a mosquito net, with a lower proportion observed in rural households (28.2%) compared to urban households (39.4%). These findings align with previous studies in sub-Saharan Africa, where net use tends to be higher in urban settings due to better availability and distribution campaigns[22]. Only a small proportion of households had mosquito netting on their windows (8.5%) or doors (3.7%), with these protective measures being more common in urban settings (window netting: 7.6% urban vs. 0.9% rural; door netting: 3.1% urban vs. 0.6% rural). There is a significant association between location, window and door netting use; urban households are more likely to use netting on doors and windows (p < 0.001), consistent with findings by Rahman *et al.,* [23] who reported greater structural prevention measures in urban dwellings.

Use of mosquito repellent spirals and sprays was widespread (82.6%) and notably more prevalent among urban households (57.0%) compared to rural households (25.6%). This highly significant association (χ² = ∼118.7, df=1; p < 0.001) agrees with other surveys indicating that urban residents rely more heavily on commercial chemical repellents due to better market access [23,24]. Conversely, the use of repellent herbs or plants was more common in rural households (14.0%; χ² = ∼59.8, df=1; p < 0.001) than in urban households (4.0%), although both remained less frequent than synthetic repellents. This trend mirrors documented traditional practices in rural communities elsewhere, where natural repellents maintain cultural significance [25].

Other mosquito control practices such as environmental hygiene, closing windows in the evening, and avoiding stagnant water were reported by only 4.8% of respondents with no statistically significant difference between urban and rural households (p = 0.08; p > 0.05), which is comparable to studies reporting low adoption rates of environmental measures in dengue-endemic areas [25].

Regarding treatment-seeking behavior upon noticing signs of dengue fever, almost all respondents (98.5%) indicated they would use modern medicine, with negligible reliance on traditional medicine (1.2%) or other responses such as uncertainty (0.38%). The observed significant difference in medication choice (urban households reported higher traditional medicine use; p = 0.008) is in line with findings from other West African settings where urban populations display slightly higher engagement with traditional healers[22].

These results highlight clear urban–rural contrasts in prevention strategies: rural households tend to rely more on personal protective behaviors and natural repellents, while urban households demonstrate greater access to structural barriers (window/door screens) and commercial chemical repellents. Despite these differences in prevention, treatment practices are overwhelmingly dominated by the use of modern medicine across both settings. This pattern of urban advantage and rural reliance on traditional methods has been reported across multiple dengue-endemic regions [3,25,26].

## 5. Conclusion

This national study conducted across all thirteen regions of Burkina Faso highlights marked disparities between urban and rural populations regarding knowledge, attitudes, and practices related to dengue fever. Despite a generally moderate level of information, urban residents demonstrate better awareness of the disease, particularly concerning transmission modes, recurrence, and the importance of preventive measures. They benefit from wider access to the media and health infrastructure, which promotes the adoption of structural prevention strategies such as installing mosquito netting on doors and windows, as well as increased use of chemical repellents. Conversely, rural populations rely more on personal protective means, notably wearing long clothing and using plant-based repellents, reflecting traditional practices and more limited resources. The near-universal reliance on modern medicine for treatment observed in both settings is a positive factor for community-based dengue control efforts, although marginal use of traditional medicine remains somewhat higher in urban areas.

These findings underline the need to intensify and diversify health education campaigns, with particular focus on rural areas where misconceptions and gaps in knowledge persist. It is also essential to implement multisectoral approaches combining mass media, mobilization of community health workers, and strengthening of local social networks to improve equitable access to information and preventive tools. Furthermore, leveraging existing practices, notably the use of mosquito nets promoted by malaria programs, could serve as an effective leverage point to reinforce dengue prevention nationally. Thus, this study provides essential data to guide national dengue control strategies aimed at reducing its health and socio-economic burden in Burkina Faso.

## Data Availability

The data underlying the findings of this study are available from the corresponding author upon reasonable request. All relevant data can be made accessible to the journal for editorial review. Data has been shared without: we have included an Excel file (S1 Dataset. Household survey data. KAP Survey) as a database in the Supporting Information

## Declarations

### Ethics approval and consent to participate

Ethical approval was obtained prior to the implementation of activities related to the protocol from the National Ethics Committee under reference number 2020-9-195, dated September 2, 2020, as well as from the Health Research Ethics Committee. All participants provided signed informed consent prior to the collection of any individual data. As all study participants were heads of households or their legal representatives and were adults, no assent or parental/guardian consent was required.

### Author’s contributions

NO and SD made substantial contributions to the study’s design, coordination of fieldwork, data analysis, and interpretation, as well as manuscript preparation. AG contributed to the design, project oversight, and substantive manuscript revision. MWG provided significant advices for the project design and implementation. HS, FT, GWN, GSS, RK, CWT, SI, and IM were responsible for data acquisition, supervision of fieldwork activities, and ensuring high-quality data collection. AG developed the electronic data collection tool used in the study. All authors revised the manuscript, approved the final submitted version, and agreed to be personally accountable for their respective contributions.

## Acknowledgements

We extend our gratitude to Burkina Faso Ministry of Health and its related institutions, national project partners, research group, regional administrative and security authorities, and project participants for their strong commitment in ensuring the success of this project.

